# Sildenafil’s Effectiveness in the Primary Coronary Slow Flow Phenomenon: A Randomized Controlled Clinical Trial

**DOI:** 10.1101/2024.01.18.24301510

**Authors:** Abbas Andishmand, Seyedmostafa Seyedhossaini, Seyedeh Mahdieh Namayandeh, Seyed Reza Mirjalili, Elnaz Adelzadeh, Amin Entezari

## Abstract

**Background:** On the one hand, the coronary slow flow phenomenon (CSFP) may cause recurrence of chest pain, prompting medical examinations and further healthcare expenses, and on the other side, it can result in myocardial infarction, ventricular arrhythmia, and sudden cardiac death.

**Objectives:** Due to the lack of agreement on the optimal treatment for CSFP, we decided to examine the effectiveness of sildenafil in this context.

**Methods:** We assessed the eligibility of 196 CSFP patients to participate in a 12-week, triple-blind, randomized, placebo-controlled study for receiving either 50 mg daily oral sildenafil or placebo. We evaluated the efficacy of sildenafil based on exercise tolerance test parameters, severity of angina, adverse effects, and major adverse cardiovascular events.

**Results:** Twenty eligible patients were randomly allocated in a 1:1 ratio to two groups. Sildenafil demonstrated significant efficacy in improving angina severity, with all recipients achieving a Class I angina severity, contrasting with a 40% attainment in the placebo group (P=0.011). Notably, Sildenafil induced statistically significant reductions in systolic and diastolic blood pressure, unlike the placebo group. Although a reduction in the QT interval favored Sildenafil (−21 millisecond vs +3 milliseconds), statistical significance was not reached (P=0.09 vs. P=0.67). Moreover, Sildenafil markedly improved Duke Treadmill Score (DTS) (P=0.005), while the placebo group showed non-significant improvement. Concurrently, the Sildenafil group exhibited significant enhancements in functional capacity (METs) and maximum heart rate during exercise testing compared to the placebo group.

**Conclusions:** We suggest that a daily low dose of sildenafil could be a valuable therapeutic option for CSFP.

**Graphical abstract:** 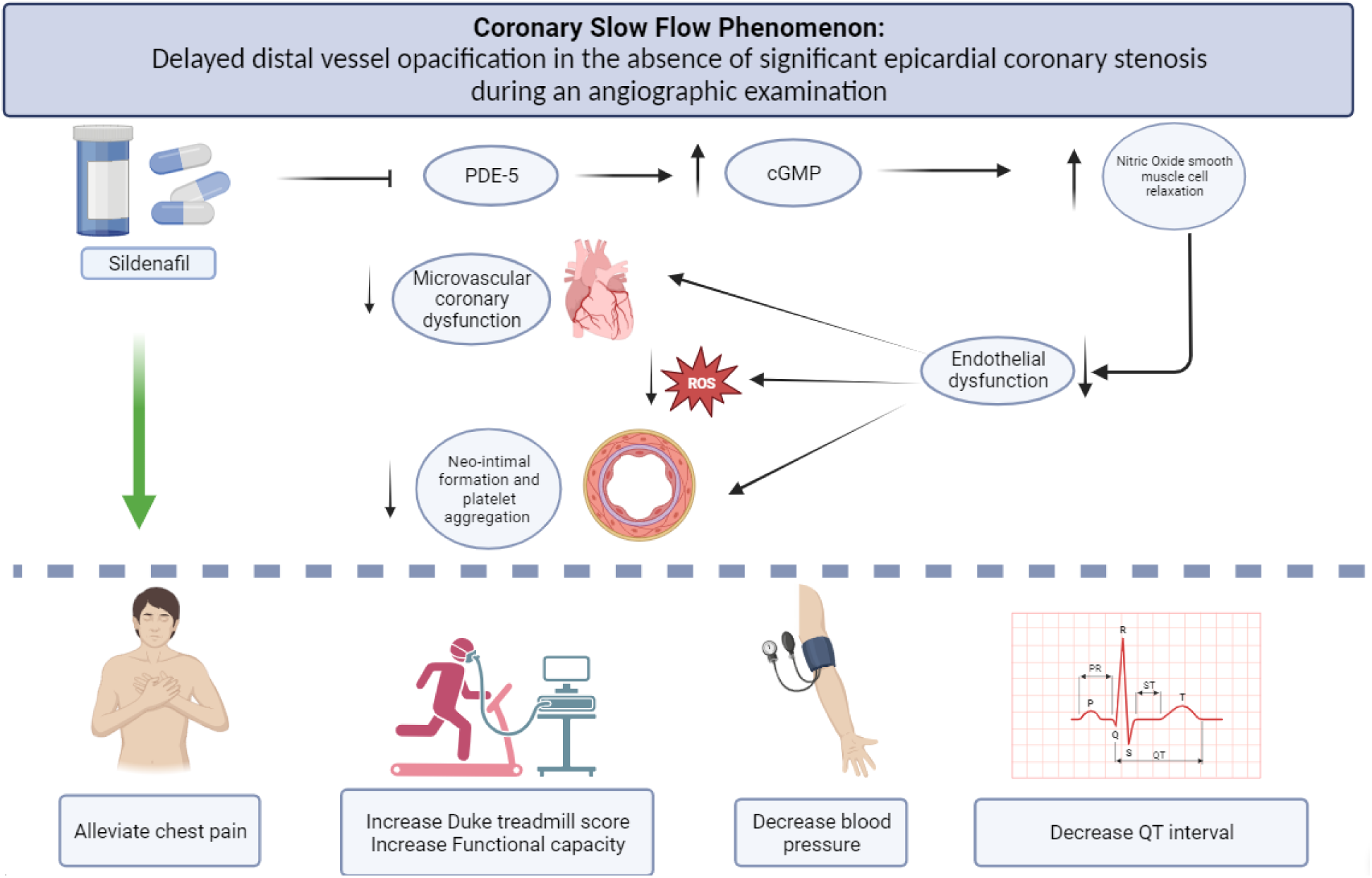

## Introduction

As the name implies, coronary slow flow phenomenon (CSFP) consists of delayed distal vessel opacification in the absence of significant epicardial coronary stenosis during an angiographic examination(1). Studies have reported different incidence rates between 1% and 7%(2), but despite its low incidence, it can be associated with recurrent chest pain and referring to medical centers, which can interfere with quality of life or even it can lead to myocardial infarction, ventricular arrhythmia, and sudden cardiac death(2). As a result, proper treatment of this problem can lead to an improvement in quality of life, a decrease in unnecessary medical evaluations and healthcare costs, and a decrease in serious cardiovascular issues.

Several types of drugs have been used as treatment for this problem, including calcium channel blockers (Diltiazem(3,4), Nicardipine(5) and mibefradil(6)), beta blockers (Nebivolol(7,8)), statins(9,10) and vasodilators (Nitroglycerin(4), Isosorbide Dinitrate(11) and Nicorandil(12)). However, there has not been any consensus regarding the optimal choice of treatment.

Knowing the pathophysiology of the problem and the drug’s pharmacodynamics is essential to choosing the optimal treatment. An important mechanism underlying the development of CSFP involves the endothelial dysfunction due to alteration of vascular tone through the reduction of nitric oxide level(13). Another hypothesis regarding CSFP is that it is linked with elevated inflammatory characteristics and oxidative stress.(14) Sildenafil hinders the activity of phosphodiesterase 5 (PDE-5) in smooth muscle cells, resulting in reduced breakdown of cyclic guanosine monophosphate(c-GMP). This, in turn, leads to elevated amounts of nitric oxide smooth muscle relaxation (15). In addition, it can exhibit antioxidant and anti-inflammatory activities(16).

Considering the above-mentioned pathophysiology of CSFP and the pharmacodynamics of sildenafil, we investigated how sildenafil affected clinical symptoms and ischemia indices in patients with CSFP in a triple-blind randomized clinical trial.

## Methods

### Trial Design

This study employed a triple-blind randomized controlled clinical trial with parallel-group design at a single center to examine the impact of sildenafil on clinical symptoms and exercise test parameters in individuals with CSFP. Twenty patients were randomly assigned in a 1:1 ratio to receive either a daily dose of sildenafil at 50 mg or a placebo. The trial has been approved by the Ethics Committee of Shahid Sadoughi University of Medical sciences, with the ethical approval number being **SSU.REC.1401.002**. In addition, all participants in the trial obtained written informed consent prior to their involvement.

In order to identify eligible patients, the Yazd Cardiovascular Disease Registry (YCDR) System and MeHR (Medical Heart Record) system reviewed all coronary angiography reports from April 2021 to September 2022 at Yazd Afshar Hospital. In order to qualify, patients were required to have a confirmed diagnosis of CSFP through coronary angiography and exhibit typical angina pectoris, with a Canadian Cardiovascular Society (CCS)(17) class greater than 1.

The diagnosis of CSFP was based on the calculation of the TIMI frame count (TFC) in each of the three territories of the coronary arteries according to Gibson’s standard criteria(18). Right coronary arteries (RCA) were marked by the first branch of the posterolateral branch artery (PLB). The point of division at the apex indicated the territory of the left anterior descending arteries (LAD) and left circumflex arteries (LCX) were demarcated by the longest obtuse marginal artery (OM). In the case of nondominant RCA, the last branch leaving the atrioventricular groove was considered, whereas in the case of dominant LCX, the last OM branch before the posterior descending (PD) branches was considered. By dividing the TIMI frame count by 1.7 for the LAD artery, the Corrected TIMI Frame Count (CTFC) was calculated, and CTFCs more than 27 in any territory were thought to be suggestive of CSFP.

Patients who satisfied any of the following criteria were barred from participating in the study: contraindication to sildenafil or use of drugs that interact with sildenafil, history of myocardial infarction or percutaneous coronary intervention (PCI) or coronary artery bypass graft (CABG), valve surgery, obstructive coronary artery disease (CAD) (stenosis above 40%), age younger than 30 years or older than 70 years, uninterpretable coronary angiography, uncontrolled hypertension, migraine headache, coronary aneurysm or ectasia, no-reflow, spasm or dissection, heart failure, valvular disease, or left ventricular ejection fraction (LVEF) less than 50%.

Among the 196 reports that were diagnosed with CSFP, only 20 individuals met the study’s inclusion and exclusion criteria and were included in the final evaluation. We used a 4-block randomization method and sealed envelope web software (https://www.sealedenvelope.com (19)) to randomly assign patients to treatment and placebo groups. To conceal the group allocation, the letters A and B were shown, and each letter was randomly assigned to either the treatment or placebo group. The number of 5 makeups of the A|B treatment group was randomized in a block of four using sealed envelope web software. The assigned treatment was written on separate papers and placed in envelopes numbered 1 to 20, which were sealed. These procedures were performed by an identified individual, and none of the other researchers were involved.

A trained nurse allocated the treatment to the patients immediately after the doctor’s visit. The nurse opened an envelope for each patient and administered the corresponding treatment from the drugs named as A and B. Both sildenafil and placebo drugs were blue round pills that were packed in coded containers to hide the identity of the prescribed drug. Finally, patients were randomly assigned in a 1:1 ratio to receive either 50mg sildenafil (Rouz darou) or a placebo once daily. It’s important to note that there were no changes to the patient’s previous medications, and only drugs containing nitroglycerin compounds were discontinued.

### Assessment, Treatment, and Follow-up

Prior to providing treatment, patients had a comprehensive baseline assessment that included interviews, physical examinations, ECGs, echocardiography and exercise testing. Clinical interview and physical examination involved recording demographic information, symptoms or clinical presentation, past medical history of CAD risk factors, medications, Body mass Index (BMI), classifying chest pain, according to CCS (17), review of the patients’ medical records, and measuring blood pressure with an Omron sphygmomanometer. Exercise tests were conducted on all patients using Track Master treadmills and the Bruce protocol (20) until symptoms became limiting. The exercise test parameters, including functional capacity measured as metabolic equivalents (METs)(21), Duke Treadmill Score (DTS)(22), maximum heart rate (HR), and HR recovery were extracted and recorded.

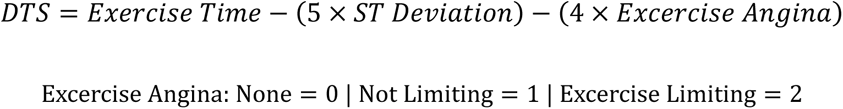

A 12-week monitoring period was conducted with monthly visits to assess medication adherence, side effects, symptoms, and CCS class. An online and telephone number allowed patients to contact a cardiology specialist 24 hours a day, and they could report any complications or new symptoms. In addition to cardiologist, an expert nurse was accountable for providing follow-up care and providing 30 pills per month during their next visit. Additionally, she kept a manual record of how patients took medicine, how many pills they took, the side effects, and their adherence. After 3 months of follow-up, all patients underwent another exercise test, and their parameters were recorded.

### Outcome Measurement

Patient outcomes were assessed by a cardiologist who was unaware of treatment allocations. The primary endpoints were changes in DTS, attained METs, improvement in clinical manifestations, and CCS class from baseline to week 12. Major adverse cardiovascular events (MACE), arrhythmia, and rehospitalization due to acute coronary syndrome (ACS) were the secondary objectives.

### Statistical Analysis

The data were analyzed using SPSS 20 software (IBM SPSS Statistics 20, Chicago, IL, USA). Quantitative data were expressed as mean and standard deviation, while qualitative data were expressed as frequency and percentage. Non-parametric tests, such as the Mann-Whitney U test and Wilcoxon Signed-Rank Test, were used to analyze quantitative variables, and the chi-square test was used to analyze qualitative data. Linear regression analysis was done to evaluate the association between TFC and Duke score. A p-value of less than 0.05 was considered statistically significant.

## Results

### Patient Characteristics

From April 2021 to September 2022, a total of 196 patients were diagnosed with CSFP through angiography. Based on the inclusion and exclusion criteria, 20 eligible patients were randomly assigned to two groups in a 1:1 ratio to receive Sildenafil or a placebo. All patients were followed-up for 12 weeks (**Figure 1**). The basic characteristics of the two groups are shown in **Table 1**, With the exception of one-minute heart rate recovery, there were no notable disparities between the intervention and placebo groups at the beginning of the study. The average age of the patients was 49 years, with 65% being male.

**Figure 1.**
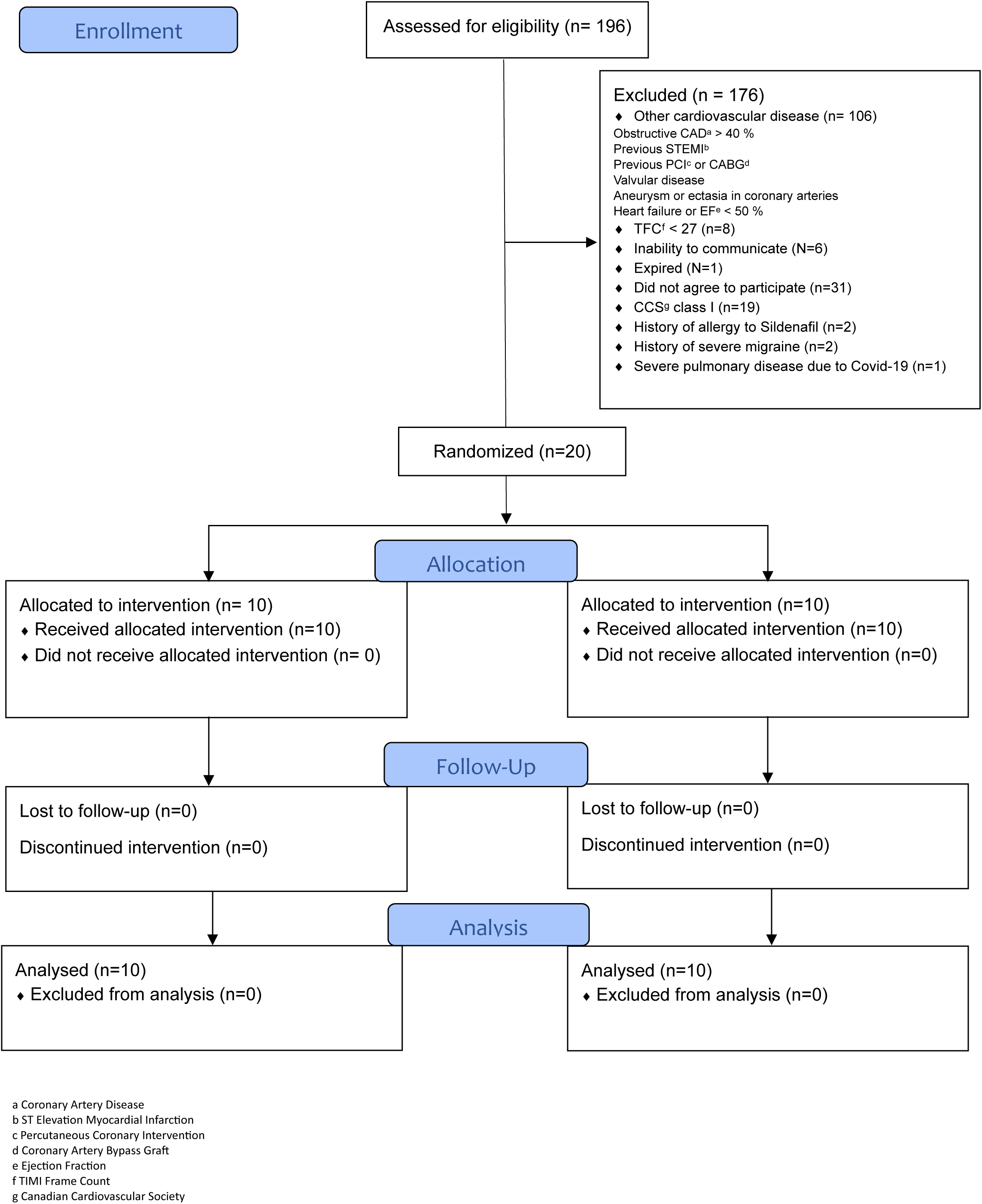
Enrollment, Randomization, Allocation and Follow-up.

**Table 1.** Baseline Characteristics of the Patients Who Underwent Randomization.

| Characteristics | Sildenafil (N=10) | Placebo (N=10) | P-value |
| --- | --- | --- | --- |
| Age (Mean ± SD) | 49.2±10.8 | 48.4±8.4 | 0.85 |
| Body Mass Index (kg/m2) (Mean ± SD) | 27.6 ± 5.3 | 26.7 ± 3.3 | 0.67 |
| Sex |  |  |  |
| Male (N (%)) | 7(53.8) | 6(46.2) | >0.99 |
| Female (N (%)) | 3(42.9) | 4(57.1) |  |
| Primary presentation |  |  |  |
| Acute coronary syndrome (N (%)) | 5(50.0) | 8(80.0) | 0.35 |
| Chronic stable angina (N (%)) | 5(50.0) | 2(20.0) |  |
| Canadian Cardiovascular Society angina class |  |  |  |
| II (N (%)) | 4 (40.0) | 6 (60) | 0.84 |
| III (N (%)) | 4 (40.0) | 2 (20.0) |  |
| IV (N (%)) | 2 (20.0) | 2 (20.0) |  |
| Risk factors |  |  |  |
| Hypertension (N (%)) | 3 (30) | 0 | 0.21 |
| Diabetes (N (%)) | 2 (20) | 0 | 0.47 |
| Hypercholesterolemia (N (%)) | 3 (30) | 3(30) | >0.99 |
| Smoking (N (%)) | 2 (20) | 4 (40) | 0.62 |
| Family history of coronary artery disease (N (%)) | 4 (40) | 7 (70) | 0.37 |
| Drug history |  |  |  |
| Nitroglycerine (N (%)) | 4 (40) | 2 (20) | 0.62 |
| Aspirin (N (%)) | 9 (90) | 8 (80) | >0.99 |
| Statin (N (%)) | 9 (90) | 6 (60) | 0.30 |
| ACEI<sup>a</sup> or ARB<sup>b</sup> (N (%)) | 3 (30) | 0 (0) | 0.21 |
| Calcium Channel Blockers (N (%)) | 1 (10) | 2 (20) | >0.99 |
| Beta Blockers (N (%)) | 7 (70) | 4 (40) | 0.37 |
| Blood pressure (mmHg) |  |  |  |
| Systolic (Mean ± SD) | 123 ± 14.8 | 127.6 ± 12.4 | 0.55 |
| Diastolic (Mean ± SD) | 74.9 ± 12.1 | 76.9 ± 8.3 | 0.67 |
| Electrocardiogram |  |  |  |
| Sinus Rhythm (N (%)) | 10 (100) | 10 (100) | >0.99 |
| Rate (Beats per minutes) (Mean ± SD) | 92.9 ± 16.4 | 82.0 ± 18.5 | 0.18 |
| ST depression (N (%)) | 4 (40) | 3 (30) | >0.99 |
| Abnormal T wave (N (%)) | 3 (30) | 6 (60) | 0.37 |
| PR interval (Mean ± SD) (Millisecond) | 136 ± 20.6 | 142 ±19.8 | 0.51 |
| QTc interval (Mean ± SD) (Millisecond) | 410.8 ± 30.7 | 426.2 ± 42.5 | 0.36 |
| Exercise Tolerance test |  |  |  |
| Positive test (N (%)) | 3 (30) | 3 (30) | >0.99 |
| Duke score (Mean ± SD) | 4.5 ± 5.5 | 4.6 ± 5.6 | 0.96 |
| METs<sup>c</sup> (Mean ± SD) | 9.92 ± 3.1 | 9.5 ± 2.1 | 0.76 |
| One-minute heart rate recovery (Mean ± SD) | 16 ± 8.4 | 25 ± 8.3 | 0.03 |
| Maximum heart rate (Mean ± SD) | 149.8 ± 17.8 | 140.7 ± 12.1 | 0.19 |
| Echocardiography |  |  |  |
| Left ventricular ejection fraction (Mean ± SD) | 53.5 ± 3.37 | 54 ± 3.1 | 0.73 |
| Angiography |  |  |  |
| Corrected TIMI frame count of LAD<sup>d</sup> (Mean ± SD) | 56.2 ± 20.7 | 42.7 ± 7.93 | 0.07 |
| TIMI frame count of LCX<sup>e</sup> (Mean ± SD) | 54 ± 14.9 | 46.6 ± 10.8 | 0.22 |
| TIMI frame count of RCX<sup>f</sup> (Mean ± SD) | 54 ± 12 | 49 ± 19.4 | 0.59 |

<sup>a</sup> Angiotensin Converting Enzyme Inhibitor<sup>b</sup> Angiotensin Receptor Blocker<sup>c</sup> Metabolic equivalents<sup>d</sup> Left Anterior descending<sup>e</sup> Left Circumflex<sup>f</sup> Right Circumflex

### Changes in severity of angina and blood pressure

After 12 weeks of treatment, it was shown that all patients who received Sildenafil experienced an improvement in the severity of angina. In fact, all of them achieved a class I CCS status, but only 40% of the patients in the placebo group exhibited the same improvement (P-value: 0.011). (**Figure 2**). Furthermore, Sildenafil treatment resulted in statistically significant reductions in both systolic and diastolic blood pressure, however, this reduction was not statistically significant with placebo treatment. (**Table 2**)

**Figure 2.**
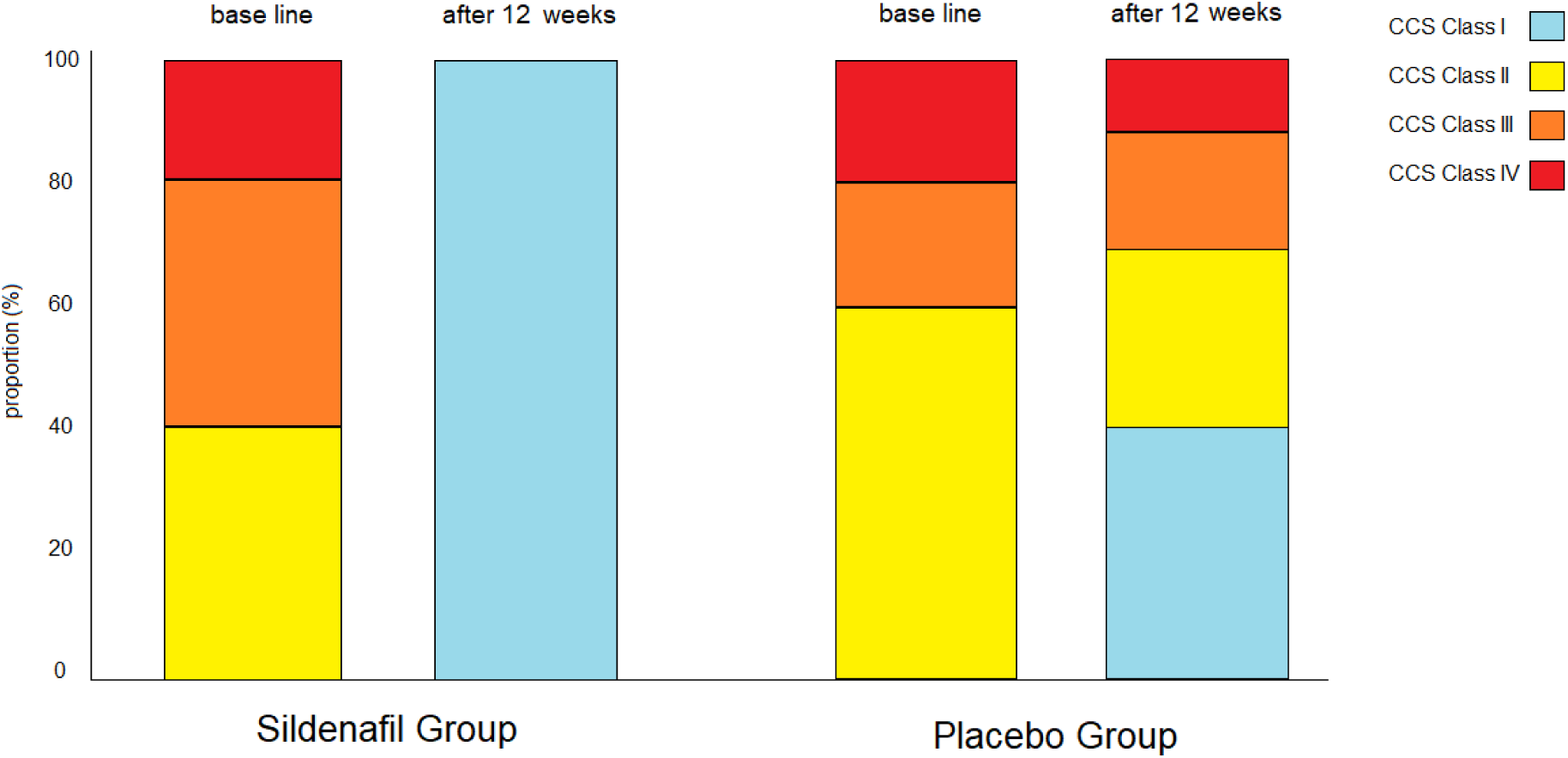
Comparison of CCS class in sildenafil group with placebo group before and 12 weeks after treatment.

**Table 2.** The investigated indices before and after treatment in sildenafil and placebo groups.

| Parameters | Sildenafil | Placebo |
| --- | --- | --- |
| **One-minute heart rate recovery** |  |  |
| Baseline (Mean ± SD) | 16 ± 8.4 | 25.1 ± 8.3 |
| After three months (Mean ± SD) | 19.2 ± 9.9 | 22.8 ± 9.8 |
| Change (Mean ± SD) | 3.2 ± 13.2 | -2.3 ± 5.3 |
| P-value | 0.45 | 0.2 |
| **Maximum heart rate** |  |  |
| Baseline (Mean ± SD) | 149 ± 17.8 | 140.7 ± 12.1 |
| After three months (Mean ± SD) | 161 ± 11.2 | 140.1 ± 10.7 |
| Change (Mean ± SD) | 12.1 ± 15.5 | 0.6 ± 12.1 |
| P-value | 0.04 | 0.87 |
| **Resting systolic blood pressure** |  |  |
| Baseline (Mean ± SD) | 123.9 ± 14.8 | 127.6 ± 12.5 |
| After three months (Mean ± SD) | 111 ± 10.8 | 117.6 ± 15 |
| Change (Mean ± SD) | -12.9 ± 8.6 | -10 ± 15 |
| P-value | 0.004 | 0.23 |
| **Resting diastolic blood pressure** |  |  |
| Baseline (Mean ± SD) | 74.9 ± 12.1 | 76.9 ± 8.3 |
| After three months (Mean ± SD) | 70.6 ± 7.1 | 78.9 ± 7.1 |
| Change (Mean ± SD) | -4.3 ± 7.5 | 2 ± 7.4 |
| P-value | 0.004 | 0.1 |
| **Corrected QT interval (Millisecond)** |  |  |
| Baseline (Mean ± SD) | 410.8 ± 30.7 | 426.2 ± 42.5 |
| After three months (Mean ± SD) | 389.8 ± 38.3 | 429.2 ± 34.7 |
| Change (Mean ± SD) | -21 ± 34.7 | 3 ± 21.8 |
| P-value | 0.09 | 0.67 |
| **Functional capacity (METs<sup>a</sup>)** |  |  |
| Baseline (Mean ± SD) | 9.9 ± 3.1 | 9.56 ± 2.1 |
| After three months (Mean ± SD) | 13.1 ± 3.3 | 9.63 ± 2.4 |
| Change (Mean ± SD) | 3.2 ± 2.8 | 0.1 ± 2.6 |
| P-value | 0.006 | 0.93 |
| **Duke score** |  |  |
| Baseline (Mean ± SD) | 4.5 ± 5.5 | 4.6 ± 5.6 |
| After three months (Mean ± SD) | 10 ± 5.3 | 6 ± 6 |
| Change (Mean ± SD) | 5.5 ± 4.8 | 1.4 ± 6 |
| P-value | 0.005 | 0.48 |

<sup>a</sup> Metabolic equivalents

### Changes in ECG and Exercise Test Parameters

Our findings indicate that Sildenafil led to a reduction in the QT interval when compared to the placebo group (−21 milliseconds versus +3 milliseconds). However, it is important to note that this difference did not reach statistical significance (P=0.09 vs P=0.67). (**Table 2**)

After 12 weeks, DTS improved significantly in the Sildenafil group, however the improvement was not significant in the placebo group (+5.5 vs +1.4; P=0.005 and P=0.48). Concurrently with these improvements, the Sildenafil group saw a significant increase in functional capacity and maximum heart rate during exercise testing, in comparison to the placebo group. (**Table 2**) Moreover, linear regression analysis revealed that higher TFC (CSFP intensity) in the Sildenafil group was associated with greater improvement in DTS (p < 0.05). There was an inverse relationship between TFC levels and DTS changes in the placebo group, where the higher the TFC level, the less improvement was observed (P < 0.05) (**Figure 3**). **Figure 4** demonstrates the resolution of ST depressions in a male participant from the sildenafil group during the third stage of exercise testing after 3 months of using sildenafil.

**Figure 3.**
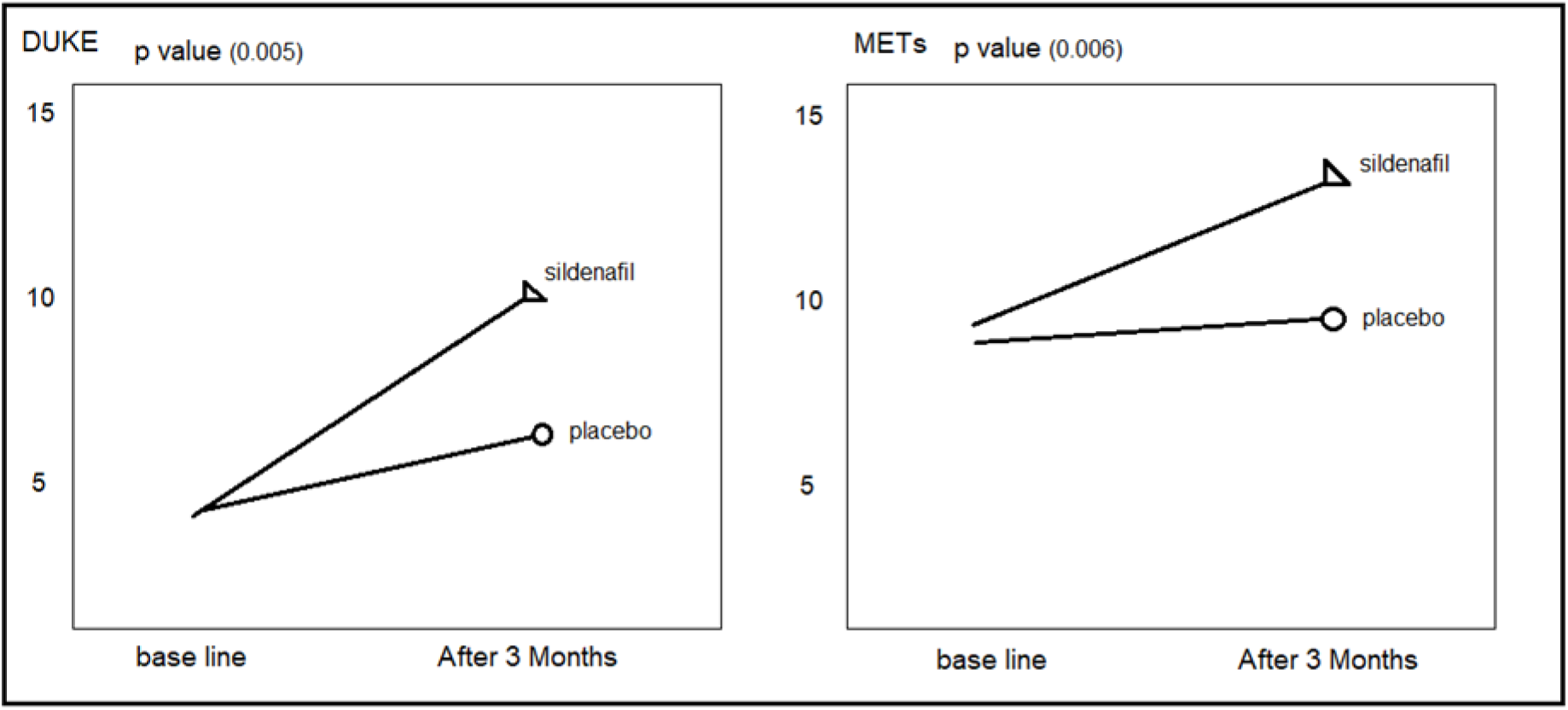
Efficacy of sildenafil versus placebo on Duke treadmill score and functional capacity (based on MET) in exercise test after 12 weeks.

**Figure 4.**
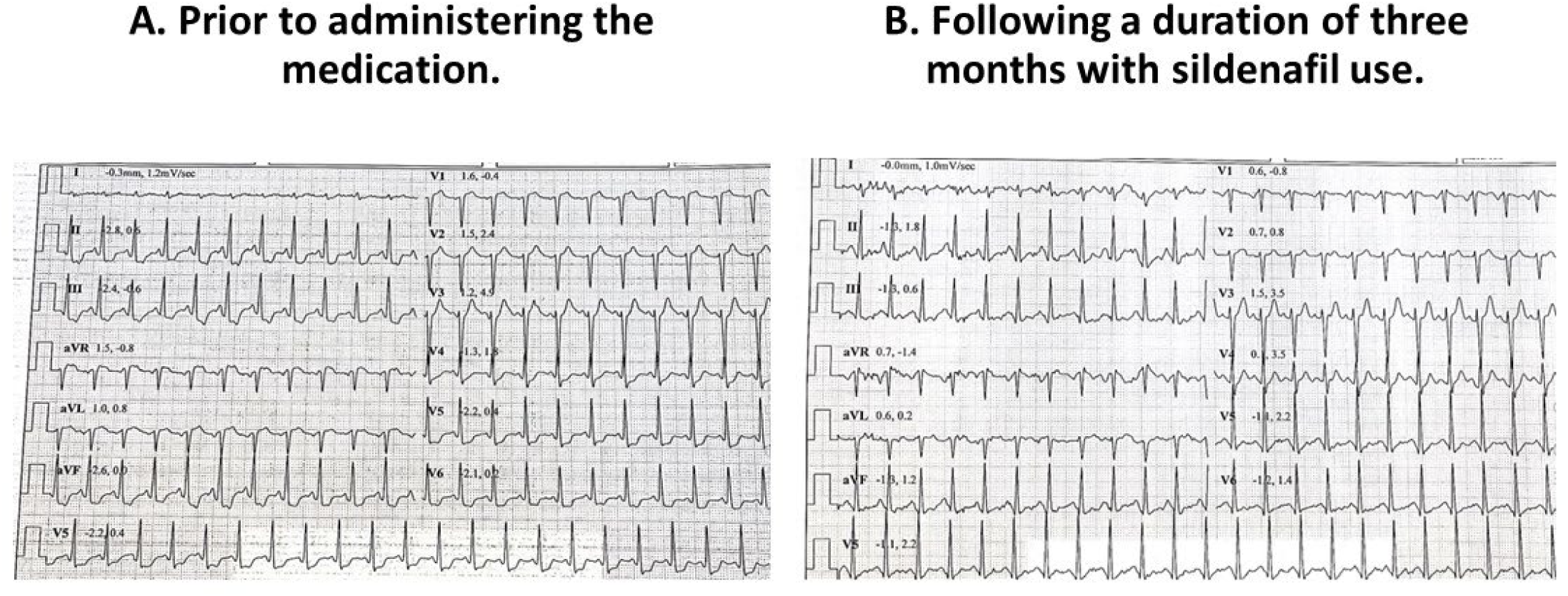
ECG recordings of a male participant who had coronary slow flow phenomenon and was enrolled in the sildenafil group.

### Adverse Events

Both groups adhered to their drug regimens completely. No cardiovascular death, arrhythmia, or myocardial infarction occurred during follow-up. Within the placebo group, a single patient was readmitted to the hospital due to acute chest discomfort, but no patients in the Sildenafil group required readmission. A tolerable headache was experienced by one patient in the Sildenafil group after taking Sildenafil for half an hour, but it completely cleared up after 2 weeks. Another patient who took Sildenafil had a transient episode of facial flushing lasting 10 minutes, which resolved spontaneously without requiring any intervention.

## Discussion

Current study is the first known investigation of the effects of PDE-5 inhibitors on CSFP. Our findings indicate that sildenafil has several benefits in CSFP, including symptoms improvement, blood pressure reduction, and enhanced exercise test parameters like as DTS, functional capacity, and maximum heart rate. Furthermore, our investigation found no notable adverse effects of the intervention, indicating that sildenafil advantages may outweigh its potential hazards in CSFP.

### Mechanism

Several theories have been proposed to elucidate the pathophysiology of CSFP, encompassing endothelial dysfunction, heightened vasomotor tone specifically within microvasculatures, platelet dysfunction, and inflammation(2). By inhibiting PDE-5, sildenafil effectively impedes the breakdown of cGMP, thereby augmenting smooth muscle relaxation in response to nitric oxide. Nitric oxide plays a pivotal role in influencing vascular tone (13) and serves as an indicator of endothelial dysfunction(23). Notably, Denardo et al. have presented compelling evidence supporting the potential of PDE-5 inhibitors to ameliorate microvascular coronary dysfunction(24). Additionally, research by Semen and colleagues has demonstrated that sildenafil contributes to a reduction in oxidative stress and modulation of fatty acid composition, implying anti-inflammatory effects(16). Furthermore, Yang et al. showcased that sildenafil effectively inhibits neointimal formation and platelet aggregation through cGMP-dependent protein kinase(25). Collectively, these findings strongly suggest that sildenafil holds promise in addressing all facets of the hypotheses posited for CSFP.

### Blood pressure

Previous studies have provided data indicating that hypertension could serve as an independent risk factor for CSFP(14,26). The present investigation found that sildenafil reduced systolic blood pressure by 13 mmHg and diastolic blood pressure by 4 mmHg. While the results were statistically significant, they may not have clinical significance in a therapeutic setting. Regardless, a decrease in blood pressure leads to a decrease in cardiac afterload, so reducing the workload on Myocytes and perhaps resulting in a decrease in the onset of symptoms.

### QT interval

Previous studies found that CSFP patients have a longer QT interval, which is a sign of an increased risk for ventricular arrhythmias and cardiovascular mortality(27,28). Despite the non-significant 0.02 second reduction in corrected QT interval, sildenafil may improve cardiac autonomic nerve tone, subsequently improving ventricular repolarization dispersion and reducing cardiac arrhythmia risk(28).

### Exercise test

In a longitudinal research conducted by Myers et al, it was shown that individuals without cardiovascular problems who died had a lower maximal heart rate compared to those who lived, following a follow-up period of 6.2 ± 3.7 years(29). Moreover, after adjusting for age, functional capacity emerged as the most influential factor in predicting mortality, both among those without cardiovascular disease and those with the condition(29). Furthermore, in a comprehensive study involving 6619 patients from two distinct cohorts, Sokari et al. identified the DTS as an indicator strongly linked to cardiovascular mortality among individuals subjected to exercise testing(30). Notably, the authors assert the superiority of METs as a predictor in this context(30). Positively influencing these predictive markers in current study, suggests a promising avenue for the potential role of sildenafil in mitigating the risks cardiovascular mortality in CSFP.

### Strength and limitation

It is important to acknowledge the following strength of the present study: The use of precise randomization and allocation concealment methods, together with the implementation of triple blinding, in this research effectively minimizes the potential for bias. Furthermore, by assessing both exercise test parameters and the symptoms shown by patients, a more focused examination of the effects of sildenafil may be conducted.

The study has several limitations. The small number of participants limits the statistical power to determine the potential adverse cardiac events and side effects of sildenafil. The follow-up did not include coronary angiography; therefore, it is unclear whether the clinical effects and improvement in ischemia observed with sildenafil are only due to improved coronary blood flow. Moreover, we were unable to compare sildenafil with other drugs that have been used in the past for CSFP treatment.

## Conclusions

Current study findings suggest that a daily low dose of sildenafil could be a valuable therapeutic option for patients with primary coronary slow flow phenomenon, by reducing chest pain, improving functional capacity, and enhancing the Duke treadmill score without any significant side effects.

## Clinical Perspectives

### What is known?

CSFP may lead to severe cardiovascular complications and substantial healthcare expenses, but there is currently no optimum therapeutic option available.

### What Is New?

We discovered that sildenafil might be an effective treatment for CSFP.

### What Is Next?

Sildenafil needs to be compared with other drugs that have been used in the past to treat CSFP in larger clinical trials.

## Data Availability

The datasets used and/or analyzed during the current study are available from the corresponding author on reasonable request.

## Abbreviations

CSFP: Coronary Slow Flow Phenomenon
DTS: Duke Treadmill Score
METs: Metabolic Equivalent
PDE-5: Phosphodiesterase 5
c-GMP: Cyclic Guanosine Monophosphate
YCDR: Yazd Cardiovascular Disease Registry
MeHR: Medical Heart Record
CCS: Canadian Cardiovascular Society
TFC: TIMI Frame Count
RCA: Right Coronary Artery
PLB: Posterolateral Branch
LAD: Left Anterior Descending
LCX: Left Circumflex Artery
OM: Obtuse Marginal
PD: Posterior Descending
CTFC: Corrected TIMI Frame Count
PCI: Percutaneous Coronary Intervention
CAD: Coronary Artery Disease
LVEF: Left Ventricular Ejection Fraction
CABG: Coronary Artery Bypass Graft
BMI: Body Mass Index
HR: Heart Rate
MACE: Major Adverse Cardiovascular Events
ACS: Acute Coronary Syndrome

## Ethics approval and consent to participate

The trial has been approved by the Ethics Committee of Shahid Sadoughi University of Medical sciences, with the ethical approval number being **SSU.REC.1401.002**. In addition, all participants in the trial obtained written informed consent prior to their involvement.

## Availability of data and materials

The datasets used and/or analyzed during the current study are available from the corresponding author upon reasonable request.

## Authors’ contributions

A.A, S.M.S, A.E were involved in the conception, design, and conduct of the study and S.M. N analyzed the data. A.E, S.R.M and E.A were involved in interpretation of the results and writing the first draft of the manuscript. All authors edited, reviewed, and approved the final version of the manuscript. A.E is the corresponding author of this work and, as such, had full access to all the data in the study and takes responsibility for the integrity of the data and the accuracy of the data analysis.

## Acknowledgments

We would like to express our gratitude to Ms. Rahmani for her invaluable efforts in collecting the data for this trial. We are also thankful to Dr. Ehsan Mirzaei, the pharmacologist who assisted us in preparing the placebo, and the cardiologists at Afshar Hospital who collaborated with us in conducting this research.

